# An Oral Heat-Inactivated Postbiotic Increases Endogenous GLP-1 and Promotes Weight Loss in Adults with Overweight or Obesity: A Randomized Placebo-Controlled Trial

**DOI:** 10.64898/2026.07.23.26358809

**Authors:** Omkara Sai Kiran Pala, Teodora Nicola, Tanmaya Madhvacharyula, Kara Siedman, Ajay Ashok, Aryan Mandot, YouFeng Yang, Amit Gaggar, Namasivayam Ambalavanan, Benjamin Bikman, Nick Norwitz, Charitharth Vivek Lal

## Abstract

**Background:** Heat-inactivated postbiotics derived from *Lactiplantibacillus plantarum* have been proposed to support weight management and metabolic health by acting on the gut–metabolic axis, including endogenous glucagon-like peptide-1 (GLP-1) signaling. resM^®^ is an orally delivered formulation combining heat-inactivated *L. plantarum RSB11* (RSB11-HI) with vitamin D3, vitamin B12, chromium picolinate, white mulberry (*Morus alba)* leaf, and fenugreek (T*rigonella foenum-graecum)* seed extracts. We tested effects of resM^®^ on body weight and a panel of metabolic outcomes in overweight or obese adults.

**Methods:** In a randomized, double-blind, placebo-controlled, parallel-group trial (NCT06911073), 80 overweight or obese adults were allocated 1:1 to oral resM^®^ or matched placebo for 8 weeks. The trial was conducted in a fully remote, direct-to-consumer setting. The primary outcome was absolute change in body weight at final scheduled weekly assessment (Week 7), corresponding to completion of the 8-week intervention. Secondary outcomes included body-mass index (BMI); a fasting metabolic laboratory panel (insulin, HbA1c, and the derived HOMA-IR); serum active GLP-1; strain-specific stool quantitative PCR for *L. plantarum* RSB11; food cravings (Food Craving Questionnaire–Trait reduced, FCQ-T-r); depressive symptoms (PHQ-9); and safety laboratories (comprehensive metabolic panel and complete blood count) with adverse-event surveillance. Between-group differences used independent-samples t-tests and baseline-adjusted ANCOVA; within-group change used paired tests.

**Results:** Body weight decreased by 2.6 kg (−3.0%) in the resM^®^ group and increased by 0.5 kg (+0.6%) on the placebo. Food craving scores fell by 15.1 points within the resM^®^ arm during the 8-week intervention (p<0.001), as did depression scores. Active GLP-1 roughly doubled among participants treated with resM^®^ and increased more than the placebo. The serum GLP-1 levels correlated with *L. plantarum RSB11* levels in the stool samples in the resM^®^ arm. Safety laboratories remained within reference ranges; gastrointestinal symptoms were more frequent with placebo than resM^®^, and no serious adverse events were reported in either group.

**Conclusions:** Over 8 weeks, resM^®^ produced clinically meaningful weight loss within-group and compared to placebo, along with improvements in food cravings, depression scores, gastrointestinal symptoms, and a 2-fold increase active GLP-1. These pilot findings support the safety and clinical efficacy of the oral post-biotic, resM^®^, for weight-management. Larger trials are needed to confirm the persistence of the clinical benefits over longer timeframes.

## 1. Introduction

GLP-1 receptor agonists have rapidly reshaped obesity care, yet roughly half of U.S. adults who start one for weight loss discontinue within a year[1], largely because of gastrointestinal intolerance, cost, and the difficulty of sustaining chronic injectable therapy. This leaves a large and growing population in need of a well-tolerated, accessible option that engages the same gut–metabolic biology without those burdens. Postbiotics that augment endogenous GLP-1 signaling are a promising candidate for that role.

Gut microbiota, microbial metabolites, intestinal barrier function, and micronutrient status all modulate enteroendocrine signaling and GLP-1 production[2–6]. Postbiotics are bioactive components derived from inactivated probiotic bacteria that retain beneficial microbial signals while offering improved stability and a more favorable safety and regulatory profile than live organisms or exogenous pharmacotherapy[7]. Heat-inactivated *L. plantarum RSB-11* (RSB11-HI) is a proprietary postbiotic that retains the bioactivity of its live counterpart; resM^®^ combines RSB11-HI with micronutrients (vitamin D3, vitamin B12, chromium picolinate) and botanical extracts (white mulberry leaf, fenugreek seed) selected for complementary roles in carbohydrate metabolism, insulin sensitivity, and appetite regulation[7–14].

Therefore, we conducted a randomized, double-blind, placebo-controlled trial in adults with overweight or obesity to evaluate the effect of oral resM^®^ on body weight (primary outcome) and on secondary metabolic and patient-reported outcomes: BMI, fasting glycemic and insulin-resistance markers, serum active GLP-1, food cravings, and depressive symptoms, together with comprehensive safety monitoring.

## 2. Methods

### 2.1 Trial design

This was a randomized, double-blind, placebo-controlled, parallel-group trial with 1:1 allocation conducted in a fully remote, direct-to-consumer setting across the United States. The trial was conducted between May 2025 and March 2026. Participants were followed for an 8-week intervention period. Participant-reported body weight and questionnaire outcomes were collected at baseline and weekly through Week 7 using an electronic study form, while fasting venous laboratory panels were obtained at baseline (Visit 1, V1) and at the end-of-study visit following completion of the intervention (Visit 2, V2). The allocation ratio and primary endpoint were fixed before analysis; no interim efficacy analysis altered the design. The analysis dataset was locked in April 2026. The trial was conducted within a superiority framework for the primary outcome; secondary and laboratory endpoints were considered exploratory. No interim analyses were performed, no formal stopping guidelines were applied, and there were no changes to the trial design, eligibility criteria, or study outcomes after trial commencement.

### 2.2 Participants and eligibility

Eligible participants were adults (≥18 years) with overweight or obesity (baseline BMI 25–40 kg/m²) who provided electronically signed informed consent. Eligibility was confirmed through a structured virtual screening process that included collection of demographic information, review of medical history, concomitant medications and dietary supplements, calculation of body mass index (BMI), and verification of all protocol-defined inclusion and exclusion criteria before randomization. Participants attended a certified diagnostic center for protocol-required fasting laboratory assessments at baseline and end of study. Height was self-reported at screening, and body weight was self-reported at baseline and weekly through Week 7 using participants’ personal household scales while wearing light clothing. BMI was calculated from the reported height and weight values. Key exclusion criteria included current use of GLP-1 receptor agonists or other prescription weight-loss medications, previous bariatric surgery, pregnancy, drug or alcohol abuse, unstable cardiovascular, hepatic, or renal disease, chronic inflammatory disorders, significant gastrointestinal or metabolic conditions affecting nutrient absorption or metabolism, concurrent participation in another clinical trial, and use of prohibited concomitant supplements. In the analyzed cohort, no participant reported pregnancy, previous bariatric surgery, weight-loss pharmacotherapy, drug dependence, or concurrent participation in another clinical trial. Alcohol consumption was ≤3 drinks/week in 77 of 80 participants (96.3%). The intervention was a self-administered oral product taken by participants at home; there were no trial sites and no clinicians or other personnel delivering the intervention, so eligibility criteria for sites and for intervention providers are not applicable.

### 2.3 Randomization and blinding

Participants who met all eligibility criteria were randomized 1:1 to resM^®^ (Group A) or matched placebo (Group B). The study was conducted using a double-blind design in which participants, study personnel, outcome assessors, data management personnel, and statisticians remained blinded to treatment allocation throughout the study. Randomization was conducted through the study’s secure electronic study management platform. After participants met all eligibility criteria and completed the required baseline assessments, the Clinical Trial Coordinator at the Contract Research Organization (CRO) performed the randomization within the platform. Assignment to the resM® or placebo group was determined by the platform according to the predefined randomization scheme and was not selectable or modifiable by the Clinical Trial Coordinator or any other study personnel. Treatment allocation remained concealed until completion of data set lock and the primary statistical analyses

### 2.4 Interventions

Participants took oral resM^®^ or matched placebo daily for 8 weeks. resM^®^ contained heat-inactivated *L. plantarum* RSB-11 (RSB11-HI) together with vitamin D3, vitamin B12, chromium picolinate, white mulberry leaf (*Morus alba*) extract, and fenugreek seed (*Trigonella foenum-graecum*) extract, with all micronutrients within established tolerable upper intake levels and all components holding GRAS status or established dietary-supplement safety profiles. The placebo contained no active ingredients and consisted of inert excipients (microcrystalline cellulose, hydroxypropyl methylcellulose, magnesium stearate, silica, carrageenan, potassium acetate, and water). Both study products were identical in appearance, packaging, and labeling to maintain blinding. No structured dietary or exercise program was prescribed. Daily compliance and any concomitant medications or supplements were recorded on the electronic form. Both the intervention and the comparator were self-administered by participants at home in a direct-to-consumer setting; no clinician or study staff delivered the intervention, and the product was shipped directly to participants. Adherence was self-reported by participants on the daily electronic form and was not verified by pill count, weight of returned product, or biochemical measure; adherence is summarized as the proportion of prescribed doses reported as taken, and the exposure-response relationship across adherence strata is reported in the exploratory analyses. Fidelity of delivery was therefore not independently assessed. With respect to concomitant care, no co-intervention was provided or withheld by the study in either group: participants continued their usual diet, physical activity, and routine medical care throughout, and concomitant medications and supplements were recorded but not restricted beyond the exclusion criteria listed above.

### 2.5 Outcomes

#### Primary outcome

Absolute change in body weight (kg) from baseline to the final scheduled weekly assessment (Week 7), corresponding to completion of the 8-week intervention.

#### Secondary and exploratory outcomes

Percent change in body weight; change in BMI; fasting metabolic panel (glucose, insulin, HbA1c) and the derived insulin-resistance indices HOMA-IR (insulin × glucose / 405, glucose in mg/dL) and QUICKI (1 / [log fasting insulin + log fasting glucose]); serum active GLP-1 and DPP-4; strain-specific stool quantitative PCR for *L. plantarum RSB11* (target-engagement marker); and food cravings measured by the Food Craving Questionnaire–Trait reduced (FCQ-T-r). Participants were instructed to undergo blood collection following a minimum 8-hour overnight fast. Prior to each scheduled laboratory visit, participants received reminders through the study application as well as follow-up telephone calls and email reminders from the study team to reinforce fasting requirements and study procedures. Laboratory analyses were performed by Quest Diagnostics.

Depressive symptoms by the 9-item Patient Health Questionnaire (PHQ-9); responder analyses (proportions achieving ≥5% or ≥10% weight loss, and laboratory normalization thresholds); and the association between craving response and weight change.

#### Safety outcomes

Comprehensive metabolic panel (liver and kidney/electrolyte markers), complete blood count (CBC), and spontaneously reported adverse events captured on the daily form over the 8-week period. Harms were defined as any untoward medical occurrence reported by a participant during the 8-week intervention period. Adverse events were collected non-systematically, by spontaneous participant report on the daily electronic form; no structured symptom checklist, solicited adverse-event list, or predefined severity grading scale was used, and events were not formally adjudicated for causality. Reported counts therefore reflect total spontaneously reported events rather than systematically ascertained incidence, and are recorded as events rather than as unique affected participants. Safety laboratory measures (metabolic panel and CBC) were, by contrast, assessed systematically in all participants at baseline and end of study.

#### Assessment schedule

Participant-reported body weight, BMI, FCQ-T-r, and PHQ-9 were assessed at baseline and weekly through Week 7. Laboratory outcomes were assessed at baseline and at the end-of-study visit following completion of the 8-week intervention.

### 2.6 Sample size

Eighty participants (40 per arm) were selected as a pilot size, sufficient to estimate the primary weight-change effect with reasonable precision and to generate variance estimates for a subsequent adequately powered trial, while remaining feasible within the study’s recruitment window and laboratory budget. Laboratory and patient-reported endpoints were considered secondary or exploratory and were not individually powered.

### 2.7 Statistical methods

Continuous outcomes are summarized as mean ± standard deviation. Within-group change from baseline was assessed with paired t-tests (with Wilcoxon signed-rank tests as a sensitivity analysis). Between-group differences were assessed with independent-samples (Welch) t-tests and, as the primary inferential model, analysis of covariance (ANCOVA) on the end-of-study value adjusted for the baseline value; adjusted between-group differences are reported with 95% confidence intervals (CIs). Standardized effect sizes (Cohen’s d / Hedges’ g) are reported for the primary outcome. Categorical responder outcomes were compared with risk ratios, odds ratios, and Fisher exact / chi-square tests. For participant reported outcomes, the final assessment was defined as the Week 7 measurement, representing the final scheduled weekly assessment during the 8-week intervention. Laboratory endpoints were analyzed using baseline and end-of-study visit measurements. Analyses used available data without imputation; the number analyzed is reported for each endpoint. Participants were analyzed in the group to which they were randomized, irrespective of adherence. The primary weight analysis included all 80 randomized participants (40 per group), all of whom completed the final follow-up visit and contributed a final weight; there were no losses to follow-up and no exclusions after randomization for the primary outcome. Secondary laboratory endpoints were analyzed on an available-case basis and included only participants with paired baseline and end-of-study samples, so the denominator varies by endpoint and is reported for each. Missing data were not imputed and no sensitivity analysis for missingness was performed; the potential for bias from available-case analysis of the laboratory endpoints is addressed in the limitations. Analyses of food cravings, PHQ-9, and the exploratory metabolic and adherence subgroups were conducted within the resM® arm only and are not between-group comparisons. Two-sided p<0.05 was considered statistically significant; given the exploratory, unadjusted nature of the secondary panel, nominal p-values are reported without multiplicity correction and are interpreted as hypothesis-generating.

### 2.8 Ethics

The trial was conducted in accordance with the Declaration of Helsinki and applicable regulatory requirements. All participants provided written informed consent prior to enrollment. No individually identifiable participant data are reported.

### 2.9 Patient and public involvement

Patients and members of the public were not involved in the design, conduct, or reporting of this trial.

## 3. Results

### 3.1 Participant flow

Participants were recruited in two enrollment phases: the first between May and June 2025 and the second between December 2025 and January 2026. All participants completed the final follow-up by March 2026. The study was conducted remotely, with participants enrolled from 20 U.S. states, including Alabama (n = 8), California (n = 14), Colorado (n = 3), Connecticut (n = 1), Florida (n = 8), Georgia (n = 7), Illinois (n = 4), Maryland (n = 1), Michigan (n = 3), Minnesota (n = 2), Missouri (n = 1), North Carolina (n = 1), New Jersey (n = 2), Nevada (n = 1), New York (n = 4), Oregon (n = 1), Pennsylvania (n = 2), Texas (n = 11), Virginia (n = 3), and Washington (n = 3). Eighty-three adults were assessed for eligibility; three were excluded before randomization due to comorbidities, concomitant medications, or clinical conditions that could compromise safety, confound the primary endpoint, or preclude protocol adherence. The remaining 80 participants were randomized 1:1 to resM® (n = 40) or matched placebo (n = 40), and all received their allocated intervention. Every randomized participant completed the follow-up period and reached the final scheduled weekly assessment (Week 7), and all were included in the primary weight analysis (40 per arm). Paired fasting metabolic laboratory values were available at both visits for the large majority of participants, and no participant was lost to follow-up for the primary endpoint **(Fig 1).**

**Figure 1.**
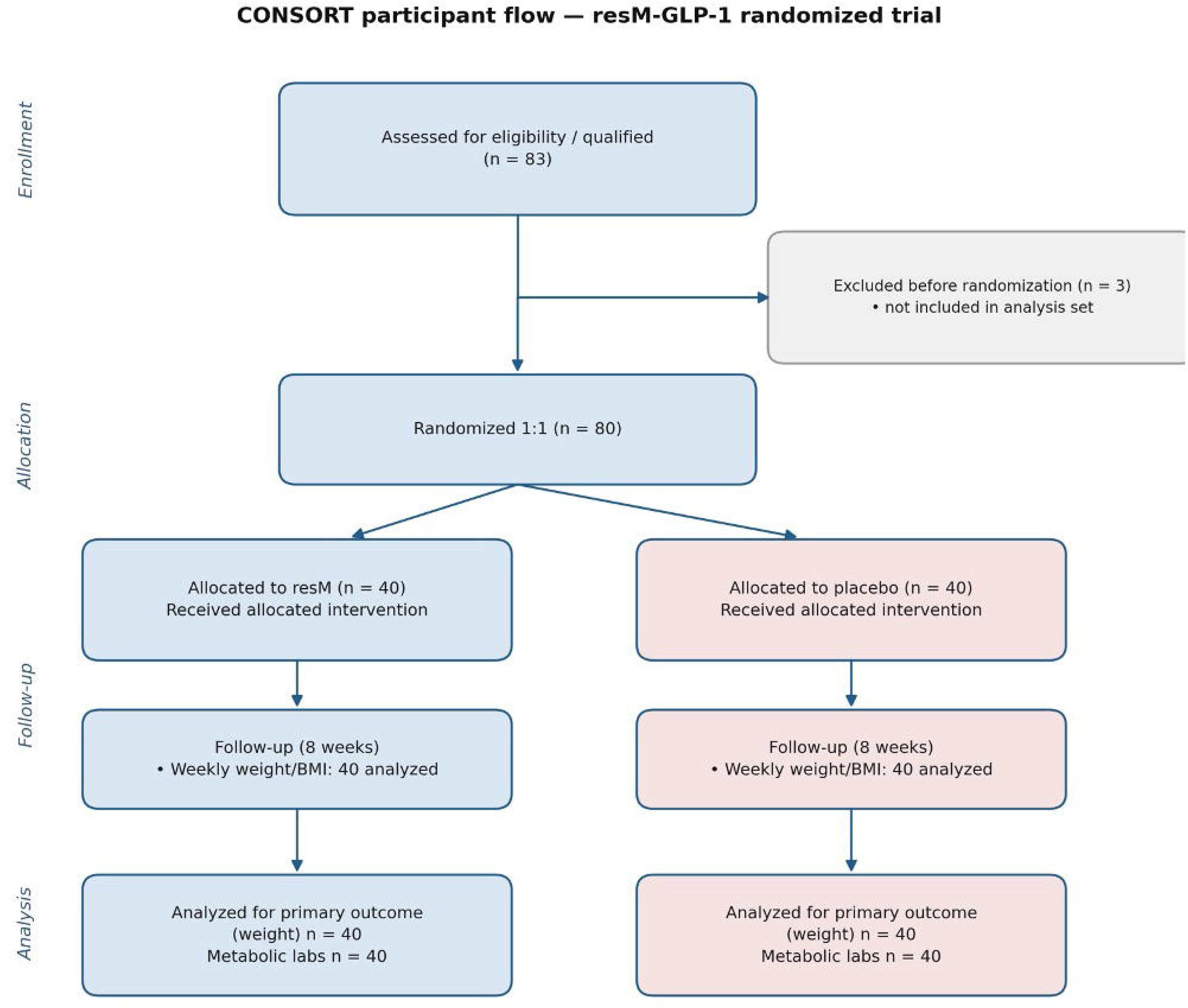
CONSORT participant-flow diagram for the resM® randomized trial, showing progression through enrollment, 1:1 allocation, follow-up, and analysis. Of 83 adults assessed for eligibility, 3 were excluded before randomization; 80 were randomized (40 resM®, 40 placebo). All randomized participants received the allocated intervention, completed follow-up, and were analyzed for the primary weight outcome (40 per arm).

### 3.2 Baseline characteristics

The two arms were well balanced at baseline (Table 1). Mean age was approximately 43 years and mean BMI approximately 31 kg/m² in both groups; the cohort was predominantly White, with Black and Asian participants represented.

**Table 1.**
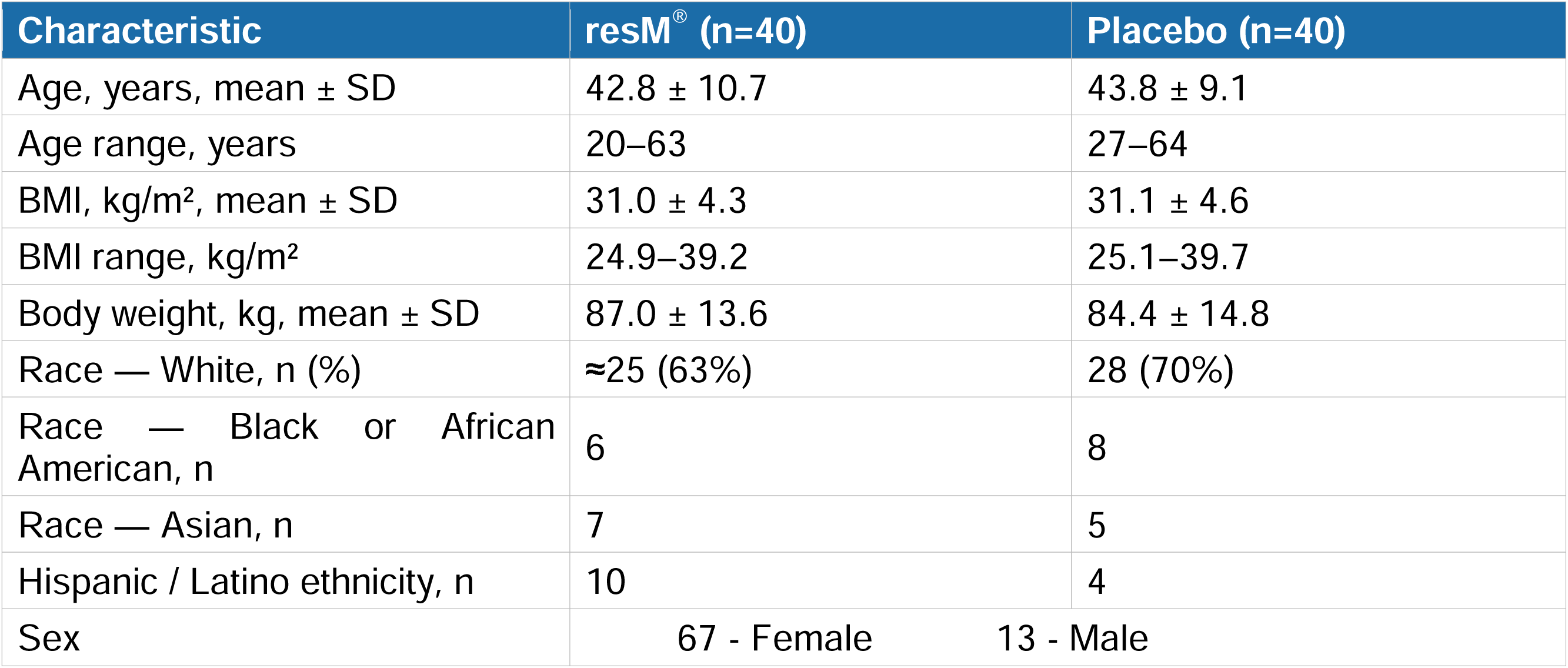
Baseline characteristics by treatment arm. Race percentages are approximate owing to multiple-race responses.

### 3.3 resM^®^ Lowers Body weight As Compared to Placebo (Primary Outcome)

resM® produced a clinically meaningful, progressive reduction in body weight within group and between group **(Fig 2)**. Specifically, by the final weekly assessment (Week 7), mean weight change was −2.6 ± 2.3 kg (−3.0%) with resM^®^ versus +0.5 ± 2.7 kg (+0.6%) with placebo. The unadjusted between-group difference was −3.11 kg (95% CI −4.22 to −1.99; p<0.001), and the baseline-adjusted (ANCOVA) difference was −3.10 kg (95% CI −4.24 to −1.97; p<0.001), corresponding to a large effect size (Cohen’s d −1.24).

**Figure 2.**
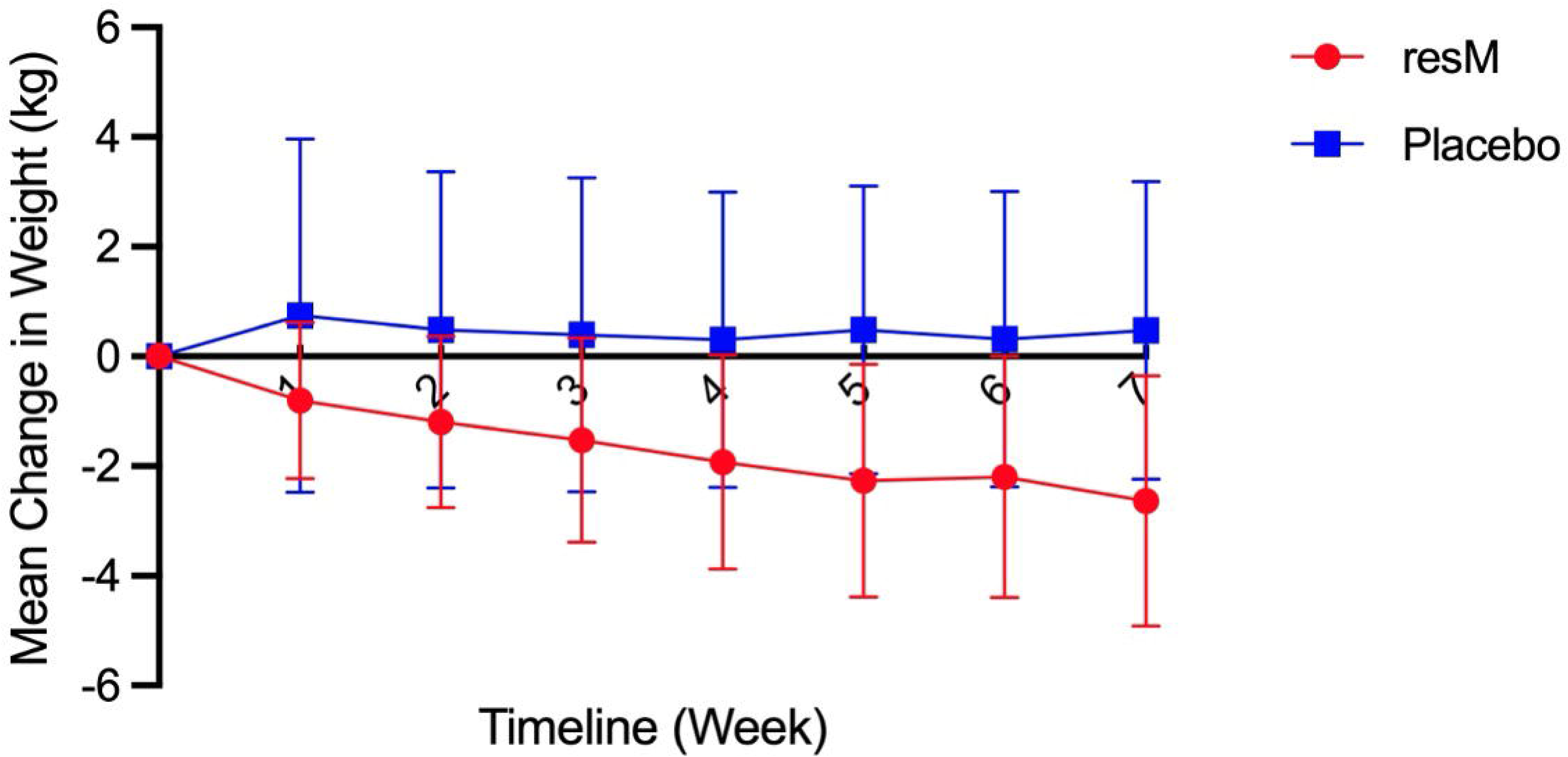
Mean change in body weight from baseline by study visit for the resM® (blue) and placebo (orange) arms, from baseline (Week 0) through Week 7. Data-label values denote the group mean at each visit. resM® showed a progressive reduction reaching −2.63 at Week 7, whereas placebo remained near or slightly above baseline throughout.

Responder analyses reinforced the primary result **(Table 2)**: 92.5% of resM^®^ participants lost weight (vs 30% placebo), 17.5% lost ≥5% of body weight (vs 2.5%), and one resM^®^ participant lost ≥10% (vs none). Conversely, 40% of placebo participants gained weight versus 2.5% with resM^®^.

**Table 2.** Primary and anthropometric (weight) outcomes at Final weekly assessment (Week 7).

| Outcome | resM<sup>®</sup> (n=40) | Placebo (n=40) |
| --- | --- | --- |
| Weight change, kg (mean ± SD) | $-2.6 \pm 2.3$ | $+0.5 \pm 2.7$ |
| Weight change, % (mean ± SD) | $-3.0 \pm 2.6$ | $+0.6 \pm 3.0$ |
| Adjusted between-group $\Delta$ (95% CI) | $-3.10$ kg ($-4.24$, $-1.97$) | $p < 0.001$; $d = -1.24$ |
| Lost any weight, n (%) | 37 (92.5) | 12 (30.0) |
| Lost $\geq 5\%$ body weight, n (%) | 7 (17.5) | 1 (2.5) |
| Lost $\geq 10\%$ body weight, n (%) | 1 (2.5) | 0 (0) |
| Gained weight, n (%) | 1 (2.5) | 16 (40.0) |

### 3.4 resM^®^ Lowers BMI

Mean body-mass index in the resM® arm decreased progressively from baseline, paralleling the primary weight result, reaching a mean change of −0.89 kg/m² by Week 7 **(Fig 3)**. The reduction was evident by Week 3 (−0.36) and sustained through the final weekly assessment, consistent with the observed weight loss.

**Figure 3.**
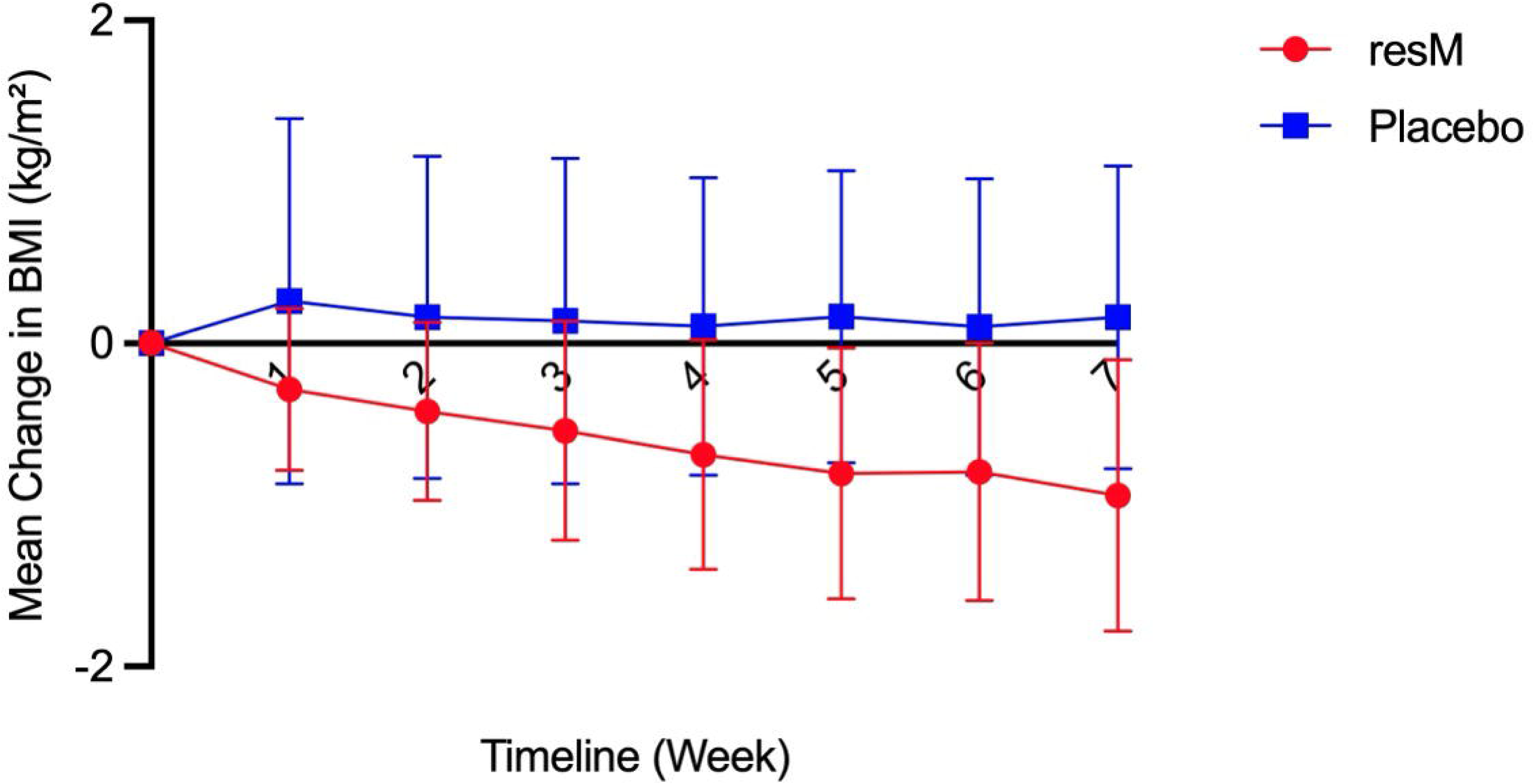
Mean change in body-mass index (BMI, kg/m²) from baseline in the resM® arm (Randomization Group A) by study visit, from baseline (Week 0) through Week 7. Points are the group mean change at each weekly visit.

### 3.5 Food cravings (FCQ-T-r)

Within the resM^®^ arm (n=40), FCQ-T-r total scores fell progressively from 51.3 at baseline to 36.3 at Week 7, a reduction of 15.1 points (95% CI −19.3 to −10.8; paired p<0.001), or approximately 29% **(Fig 4)**. The reduction was evident by Week 1 and sustained through Week 7. Greater craving reduction was associated with greater weight loss (participants with ≥10-point FCQ reduction lost −2.9 kg on average vs −2.1 kg for smaller reductions; Pearson r=0.32, p=0.045). Craving data were analyzed within the resM^®^ arm and are not presented as a between-group comparison.

**Figure 4.**
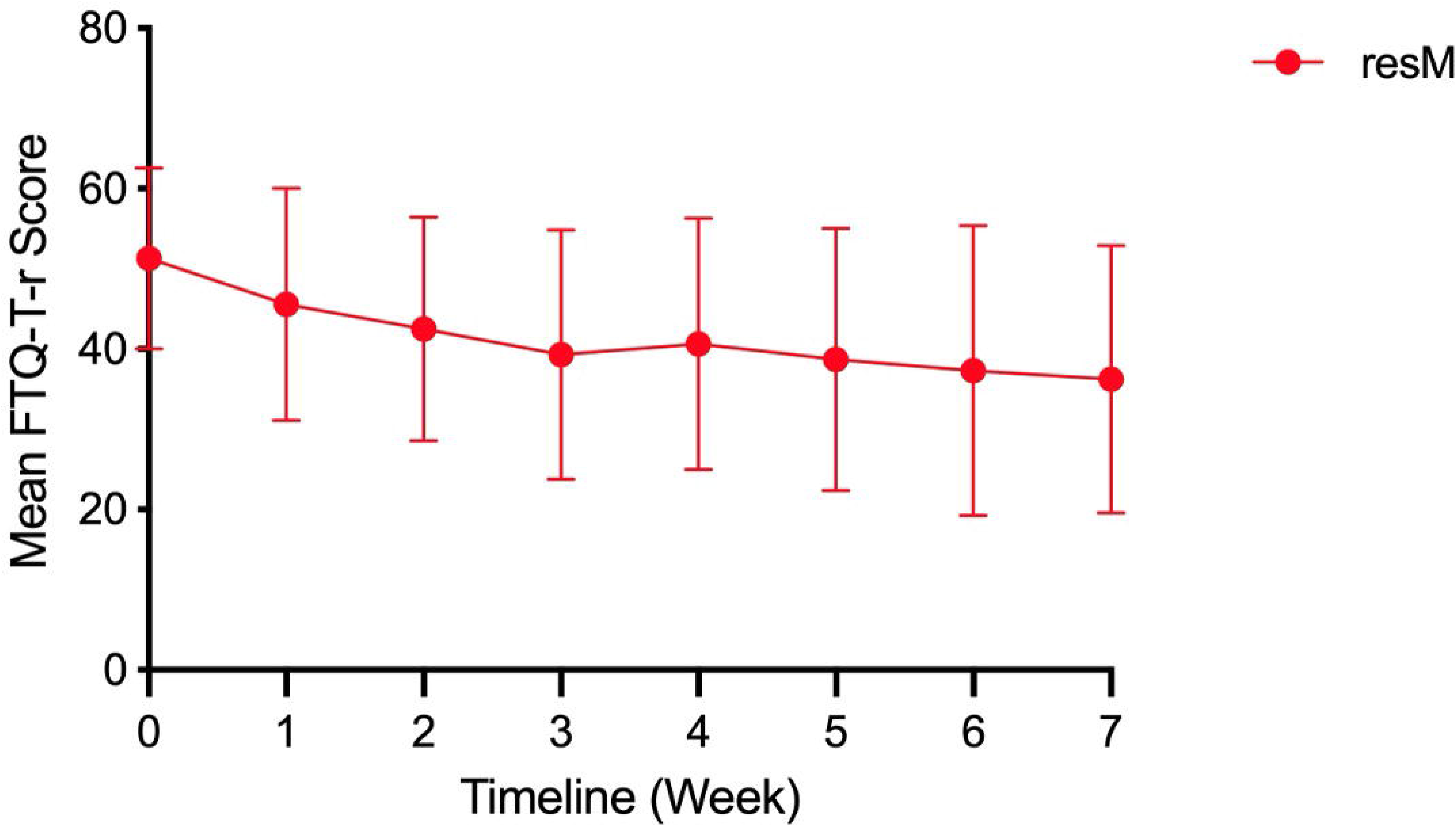
Food-craving trajectory (FCQ-T-r total score) in the resM® arm from baseline (Week 0) through Week 7. Points are the arm mean at each visit. Craving data were analyzed within the resM® arm and are not presented as a between-group comparison.

### 3.6 Active GLP-1 levels and Strain-specific stool PCR (target engagement)

Serum active GLP-1 (7-36 amide) more than doubled over 8 weeks in the resM^®^-treated participants (mean ∼6.5 to ∼15 pg/mL; p<0.001), whereas GLP-1 levels in the placebo arm were essentially unchanged (∼7.5 to ∼8.5 pg/mL) **(Fig 5A)**. At the end of study, active GLP-1 in the resM^®^ arm was significantly higher than its own baseline and greater than the placebo baseline and end-of-study values (p<0.001, each). Post-prandial GLP-1 dynamics were assessed.

**Figure 5.**
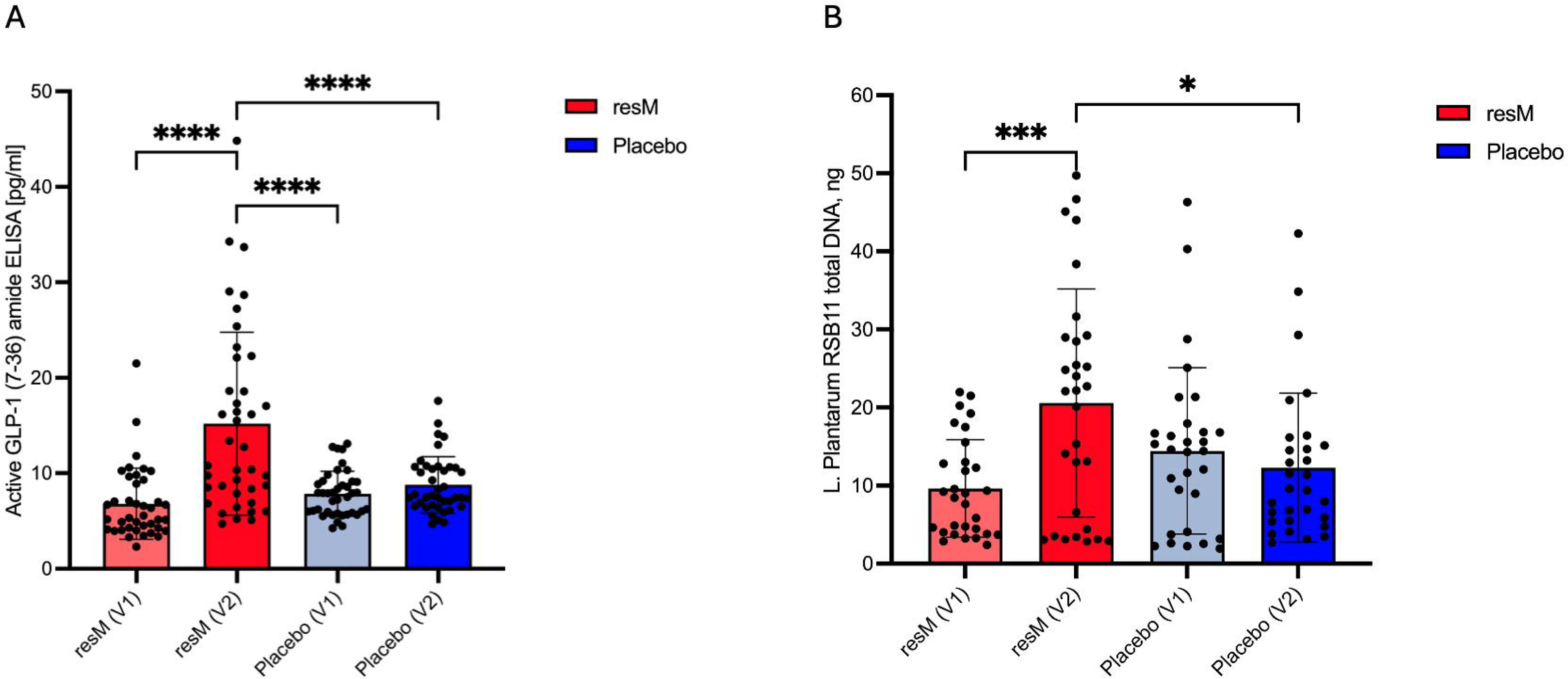
**(A)** Serum active GLP-1 (7–36 amide, pg/mL) by group at baseline and end of study (all 80 participants). Bars show mean ± SD. Active GLP-1 increased more than 2-fold in the resM® arm, with the resM® end-of-study value significantly greater than its own baseline, the placebo baseline, and the placebo end-of-study value (****p < 0.0001 for each comparison). **(B).** Strain-specific stool PCR for L. plantarum RSB11 by group at baseline and end of study (n = 60). Bars show mean ± SD. RSB11 stool DNA rose ≈ 2.5-fold in the resM® arm (***p < 0.001 vs resM® baseline) and was higher than placebo at end of study (*p < 0.05), confirming target engagement.

To verify that the administered postbiotic strain reached the gastrointestinal tract, strain-specific quantitative PCR for *L. plantarum RSB11* was performed on stool from a subset of 60 participants at baseline and end of study. In the resM^®^ arm, RSB11 stool DNA increased approximately 2.2-fold over 8 weeks (mean ∼9.5 to ∼20.5 ng; within-arm p<0.001) interestingly, by approximately the same proportion as active GLP-1 in the serum, whereas the placebo arm did not exhibit any significant increase. At end of study, RSB11 DNA in the resM^®^ arm was significantly higher than in the placebo arm (p<0.05) **(Fig 5B)**. This data provides direct evidence of target engagement, detection of the strain-specific genomic signal in stool, distinguishing the active arm.

### 3.7 Glycemic and insulin-resistance markers

Within the resM® arm, all three fasting metabolic markers moved in a favorable direction from baseline (V1) to end of study (V2), although none reached statistical significance in this pilot sample: fasting insulin declined from ≈ 15.5 to ≈ 13.8 µIU/mL (p = 0.37) **(Fig 6A)**, HbA1c from ≈ 5.55% to ≈ 5.45% (p = 0.11) **(Fig 6B)**, and HOMA-IR from ≈ 3.70 to ≈ 3.60 (p = 0.49) **(Fig 6C).** Thus, while not statistically significant, the within-arm changes reflected a consistent favorable trend toward improved fasting glycemia and insulin sensitivity with resM®.

**Figure 6.**
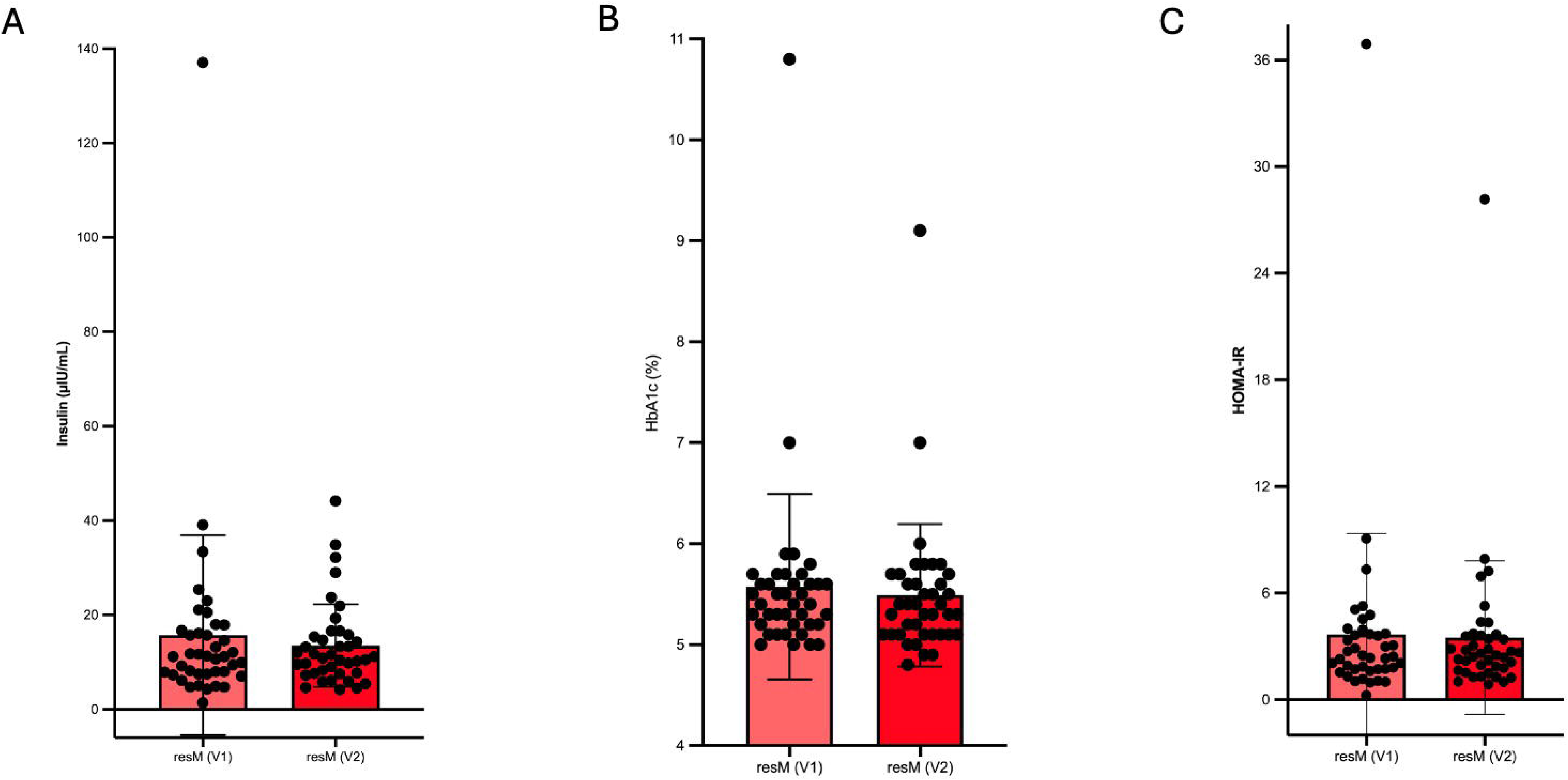
Fasting metabolic markers in the resM® arm (Group A) at baseline (V1) and end of study (V2): **(A)** fasting insulin (µIU/mL), **(B)** HbA1c (%), **(C)** HOMA-IR. Bars show mean ± SD; within-group paired p-values are shown on each panel. Although none of the within-arm changes reached significance, all three markers trended in a favorable direction (lower insulin, HbA1c, and HOMA-IR at end of study).

### 3.8 Exploratory subgroup and responder analyses

Pre-specified weight and BMI trajectories were examined by subgroup. Weight loss with resM^®^ was evident across all age bands but was largest in participants aged 18 to 39 years, reaching roughly 1.8 to 2.2 kg of mean loss by Week 7, and was smallest in the 55 to 64 age band **(Fig 7A)**; BMI reductions followed the same age gradient **(Fig 7B)**. By sex, both groups lost weight, with a larger absolute reduction in men (about 1.6 kg) than women (about 1.0 kg), consistent with higher baseline body weight in men **(Fig 7C)**. Weight loss was present across baseline BMI categories.

**Figure 7.**
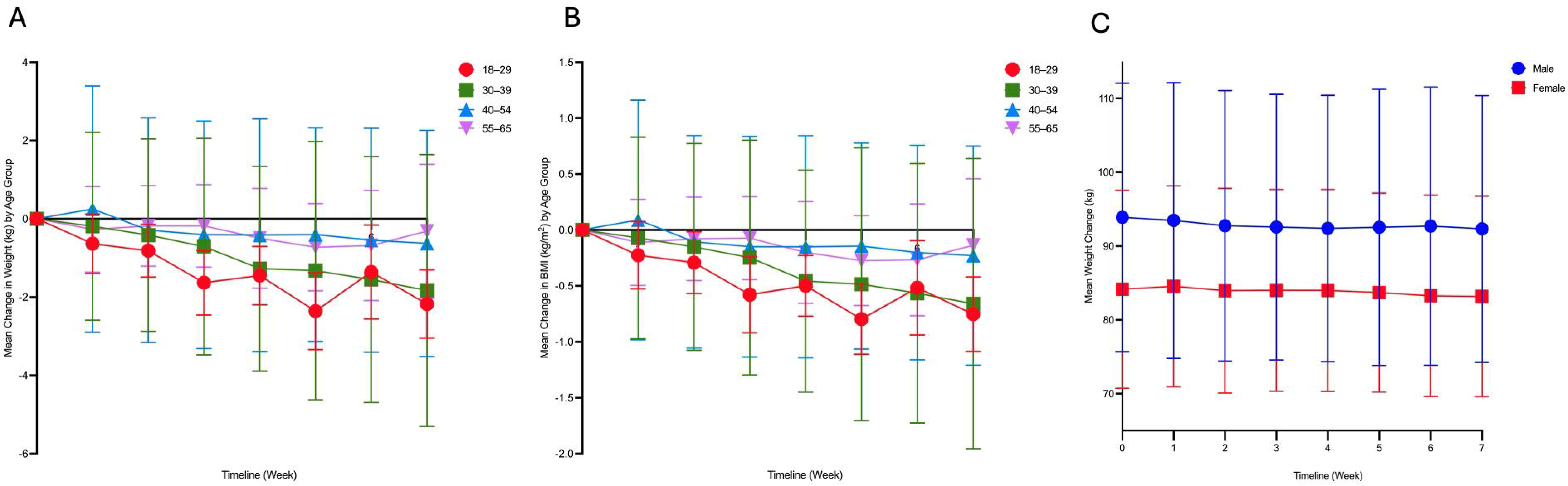
Exploratory anthropometric subgroup analyses in the resM® arm, from baseline (Week 0) through Week 7. **(A)** mean weight change (kg) by age group; **(B)** mean BMI change by age group; **(C)** mean body weight by sex. Age groups: 18–29, 30–39, 40–54, and 55–64 years. Subgroup cells were small; analyses are descriptive and hypothesis-generating.

### 3.9 Exploratory metabolic and adherence subgroup analyses

Metabolic and adherence subgroups were explored within the resM^®^ arm. Reductions in HbA1c were concentrated in participants entering with an elevated HbA1c (≥6.5% flag), with little change in those with normal or prediabetes-range values, indicating that any glycemic movement was driven by the highest-baseline stratum **(Fig 8A)**. Weight loss was greater in high-adherence participants (≥85% compliance; about 1.1 kg) than in lower-adherence participants (<85%; about 0.5 kg), supporting an exposure-response relationship **(Fig 8B)**. These exploratory, within-arm, baseline-stratified analyses are hypothesis-generating and were not controlled for multiplicity.

**Figure 8.**
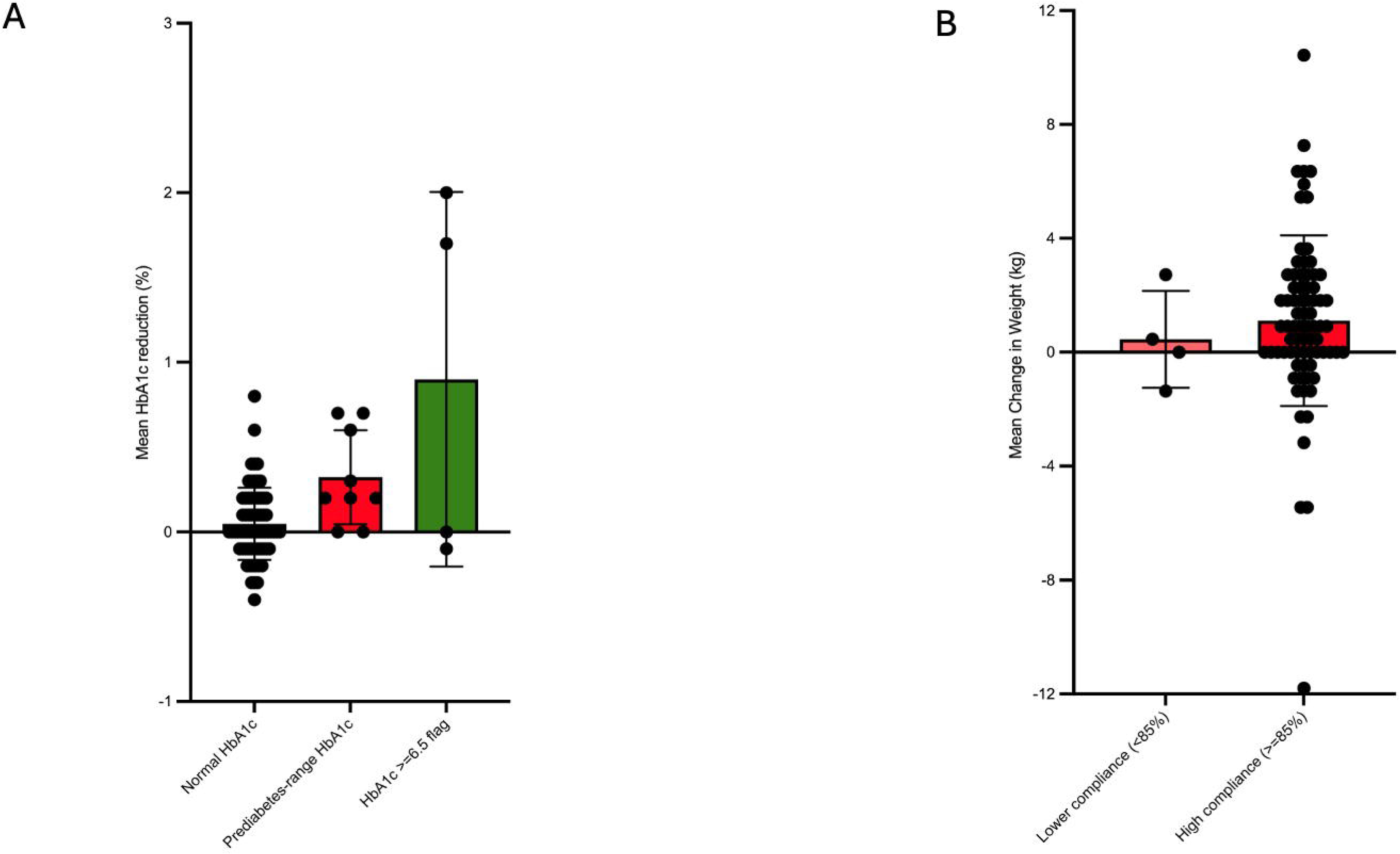
Exploratory metabolic and adherence subgroup analyses (resM® arm). **(A)** mean HbA1c change by baseline HbA1c category (normal, prediabetes-range, ≥6.5% flag); center: mean HOMA-IR change by baseline insulin-resistance status (higher vs lower baseline HOMA-IR); **(B)** mean weight loss by compliance group (≥85% vs <85%). These within-arm, baseline-stratified analyses are hypothesis-generating and were not adjusted for multiplicity.

### 3.10 Depressive symptoms (PHQ-9)

Depressive-symptom severity shifted toward milder categories in the resM® arm from baseline to the final weekly assessment (Week 7) **(Fig 9).** The proportion of participants classified as minimal severity rose from 50.0% to 67.5%, the mild category fell from 30.0% to 22.5%, and the moderate category fell from 20.0% to 10.0%. Consistent with this shift, mean PHQ-9 improved within the resM® arm (−2.05, paired p < 0.001); the baseline-adjusted between-group difference did not reach significance.

**Figure 9.**
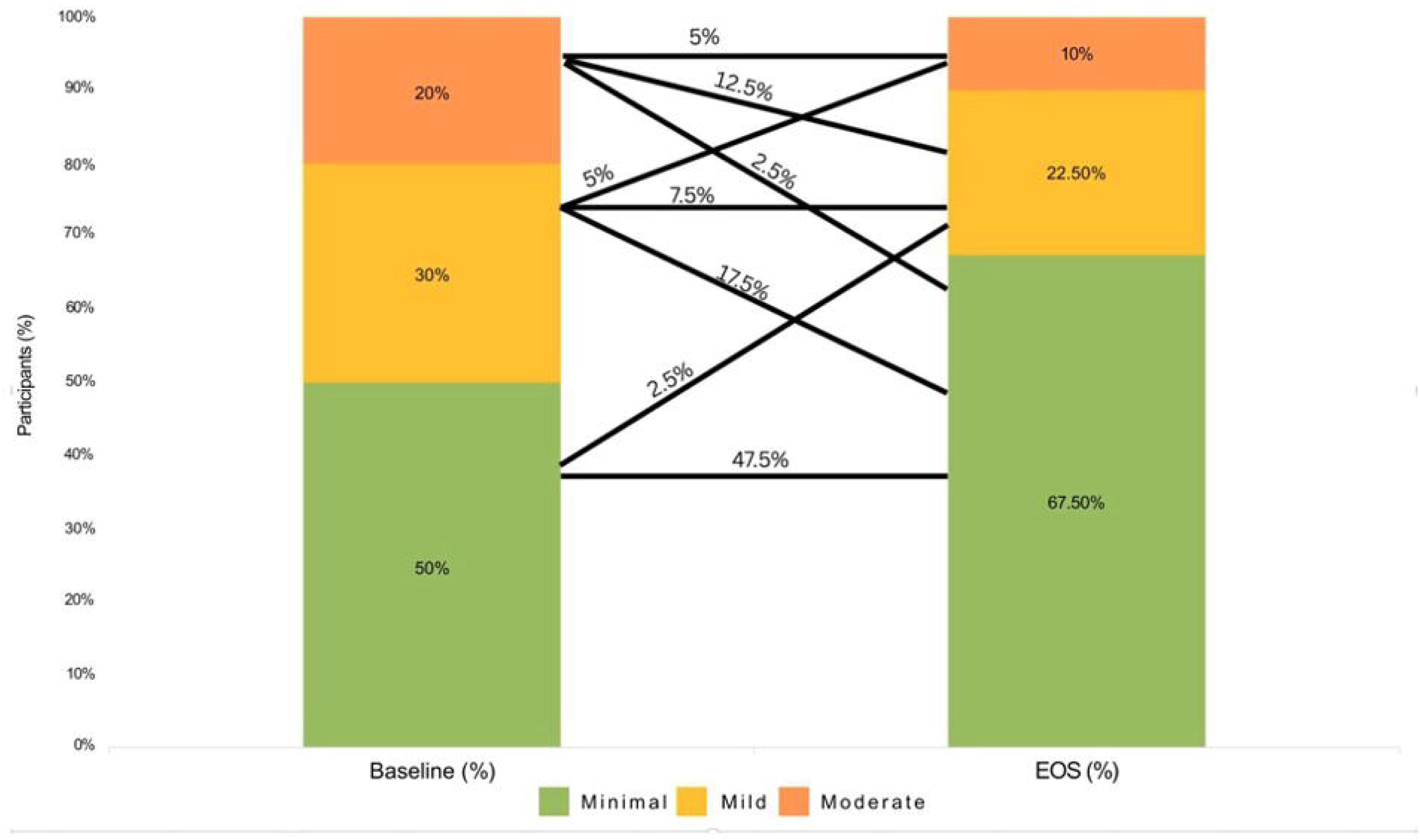
PHQ-9 depression-severity distribution in the resM® arm at baseline versus the final weekly assessment (Week 7). Stacked bars show the percentage of participants in each severity category. The distribution shifted toward the minimal category over the trial (minimal 50.0% → 67.5%; mild 30.0% → 22.5%; moderate 20.0% → 10.0%).

### 3.11 Safety and tolerability (harms)

No serious adverse events were reported. Safety laboratories including comprehensive metabolic panel (liver enzymes, bilirubin, albumin/globulin, renal markers, electrolytes) and CBC indices remained within reference ranges in both arms, with no clinically meaningful between-group differences; a small platelet-count divergence (resM^®^−10.7 vs placebo +7.6 ×10□/L; adjusted p=0.77) occurred entirely within the normal range and was not associated with any clinical event.

Spontaneously reported adverse events were predominantly gastrointestinal and were more frequent with placebo than with resM^®^ **(Table 3)**. Reported event counts over 8 weeks (resM^®^ vs placebo) were: bloating 4 vs 35, gas/flatulence 2 vs 11, abdominal pain 0 vs 9, constipation 1 vs 5, nausea 1 vs 3, and vomiting 0 vs 1; diarrhea was more frequent with resM^®^ (4 vs 0). Non-gastrointestinal reports were infrequent in both arms (headache 1 vs 3; sleep-related 1 vs 1). Counts reflect total reported events, not unique participants, and some events clustered within individuals.

**Table 3.** Adverse-event reports by group (total event counts over 8 weeks). No serious adverse events were reported in either arm.

| AE category | resM® events | Placebo events | Serious |
| --- | --- | --- | --- |
| Bloating | 4 | 35 | 0 |
| Gas / flatulence | 2 | 11 | 0 |
| Abdominal / stomach pain | 0 | 9 | 0 |
| Constipation | 1 | 5 | 0 |
| Nausea | 1 | 3 | 0 |
| Diarrhea | 4 | 0 | 0 |
| Vomiting | 0 | 1 | 0 |
| Headache | 1 | 3 | 0 |
| Sleep-related | 1 | 1 | 0 |

### 3.12 Safety laboratories within the resM® arm

Safety laboratories remained within reference ranges through end of study. Within the resM® arm, platelet count decreased modestly from ≈ 276 to ≈ 266 × 10□/L (within-arm p = 0.11) but stayed well within normal limits; hematocrit (≈ 42.7% to ≈ 42.3%; p = 0.29), AST (≈ 19 to ≈ 17.8 U/L; p = 0.18), creatinine (≈ 0.81 to ≈ 0.82 mg/dL; p = 0.56), and ALT (≈ 21.3 to ≈ 19.4 U/L; p = 0.19) were essentially unchanged **(Fig 10).** No safety-laboratory change was clinically meaningful, and no serious adverse events were reported in either arm.

**Figure 10.**
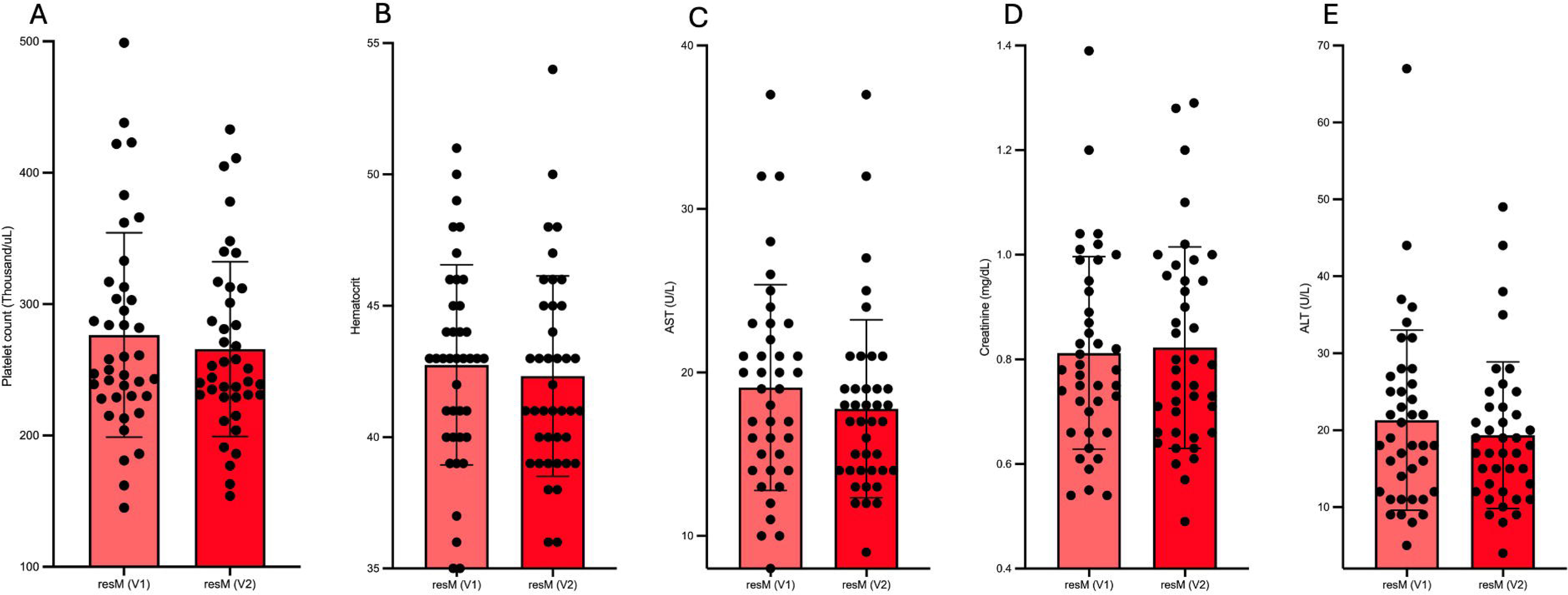
Safety laboratories in the resM® arm at baseline (V1) and end of study (V2): **(A)** platelet count (×10□/L), **(B)** hematocrit, **(C)** AST (U/L), (**D)** creatinine (mg/dL), **(E)** ALT (U/L). Bars show mean ± SD; within-group paired p-values are shown on each panel. All values remained within reference ranges; the small platelet-count change occurred entirely within the normal range.

## 4. Discussion

In this randomized, double-blind, placebo-controlled trial, 8 weeks of oral resM^®^ produced a clinically meaningful reduction in body weight, approximately 3.1 kg (about 3% of body weight) greater than placebo, a large effect accompanied by a high responder rate, a sustained within-arm reduction in food cravings, a greater than two-fold increase in serum active GLP-1, and favorable gastrointestinal tolerability.

The weight and craving results are internally consistent and biologically coherent: craving reduction tracked with weight loss, the GLP-1–satiety axis offers a plausible appetite-mediated mechanism, and the favorable gastrointestinal profile (notably less bloating, gas, and abdominal pain than placebo) is consistent with a postbiotic acting on the gut.

That said, and despite the clear clinical promise of the resM®, with emphasis on the positive primary outcome (weight change), and concordant improvements in food cravings, one apparently paradoxical finding deserves unpacking: the seemingly lower HOMA-IR and insulin resistance indices in the placebo group initially appear discordant with the approximately 2-fold increase in active GLP-1 and the statistically significant weight loss observed with resM®.

First, it is important to emphasize that this pilot study was not powered to detect differences in these secondary metabolic endpoints. Moreover, the absolute changes in fasting glucose and insulin, the variables from which HOMA-IR is derived, were small and individually non-significant, raising the possibility that the observed between-group difference reflects statistical noise rather than a true biological effect.

Nevertheless, there are plausible physiological explanations that merit consideration. Endogenous GLP-1 acts as an incretin hormone, stimulating glucose-dependent insulin secretion in the postprandial state. However, this does not necessarily translate into higher fasting insulin concentrations. Indeed, treatment with GLP-1 receptor agonists is generally associated with reductions in insulin exposure over time, as reflected by lower insulin area under the curve, owing to reduced energy intake, slower gastric emptying, lower effective glycemic exposure, weight loss, and weight-loss associated improvements in insulin sensitivity[15].

Importantly, the present study assessed glucose and insulin only in the fasting state and did not include standardized postprandial measurements. Consequently, we captured only a limited snapshot of metabolic physiology. If resM® exerts analogous effects to enhanced endogenous GLP-1 signaling including increased pre-meal satiety via hypothalamic pathways, delayed gastric emptying, and reduced effective glycemic load participants would be expected to consume less food and experience lower overall postprandial glucose and insulin excursions. Such changes could reduce cumulative insulin exposure throughout the day, consistent with the observed weight loss, while producing variable changes in fasting glucose and insulin over a short-term (8 week) trial.

In summary, the apparent discordance between HOMA-IR and the favorable changes in GLP-1 and body weight may reflect both the limited statistical power of this pilot study and the exclusive reliance on fasting metabolic measurements. Larger, adequately powered trials incorporating standardized meal challenges with serial postprandial glucose and insulin measurements will be important for more fully characterizing the metabolic effects of resM®.

Additionally, the observed improvements in gastrointestinal symptoms and mood warrant further discussion, as participants receiving resM® experienced improvements in both domains. Although the study was not designed to determine causality, these findings are biologically plausible and may reflect interconnected mechanisms operating along the gut–brain axis.

Psychological stress is known to influence gastrointestinal physiology through bidirectional neural, endocrine, and immune pathways, including vagal signaling[16]. For example, psychological stress impairs gut health and function through top-down control via the vagus nerve, altering secretions from Brunner’s glands that can ultimately lead to microbiome dysbiosis, increased intestinal permeability, and symptoms such as bloating[17]. Therefore, any intervention that improves mood and gastrointestinal symptoms could have a positive reinforcing effect over time, with improved gut health improving mood, and improved mood and mental health improving gut health.

These observations raise the possibility that gut-targeted postbiotics such as resM® may have applications beyond weight management. In particular, they suggest a potential role within the emerging field of metabolic psychiatry, which examines how metabolic interventions including those targeting the gut microbiome may influence mental health outcomes. Although exploratory, this represents an intriguing avenue for future investigation.

### 4.1 Limitations

This trial has several limitations. With respect to potential bias, the trial was conducted in a fully remote, direct-to-consumer setting, and the primary outcome relied on participant self-measured body weight recorded on an electronic form rather than on a calibrated study scale; scale make, calibration status, and measurement resolution were not standardized across participants, and adherence was self-reported and not independently verified. Additionally, because the study was conducted remotely, participant eligibility relied primarily on self-reported medical history and protocol-defined eligibility criteria rather than standardized in-person physician examinations, and stable comorbidities not meeting exclusion criteria may have been present. Although participants were instructed to fast for at least 8 hours before blood collection and received reminders through the study application, telephone calls, and email follow-ups before scheduled laboratory visits, fasting status relied on participant adherence and was not independently verified. Secondary laboratory endpoints were analyzed on an available-case basis without imputation, so differential availability of paired samples could introduce bias. Food craving, PHQ-9, and the metabolic and adherence subgroup analyses were conducted within the resM® arm only and lack a placebo comparator, so within-arm changes in these endpoints cannot be attributed to the intervention. With respect to imprecision, the trial used a fixed pilot sample of 80 participants and was powered only for the primary weight outcome; the secondary metabolic endpoints were not individually powered, and their confidence intervals are correspondingly wide, so null findings in this panel should not be read as evidence of absence of effect. With respect to multiplicity, a large number of secondary and exploratory endpoints, responder thresholds, and baseline-stratified subgroups were examined without correction for multiple comparisons; the reported nominal p-values for these endpoints are therefore hypothesis-generating and carry an elevated risk of type I error. With respect to generalisability, participants were self-selected volunteers recruited in a direct-to-consumer setting in the United States, were predominantly female and predominantly White, and were followed for only 8 weeks; the findings may not generalize to more diverse populations, to individuals with significant comorbidity or established metabolic disease, to clinically supervised or outpatient settings, or to longer periods of use, and the durability of the observed weight loss beyond 8 weeks is unknown.

## 5. Conclusions

Over the 8-week intervention, the heat-inactivated *L. plantarum RSB-11* postbiotic formulation resM^®^ produced clinically meaningful, placebo-controlled weight loss in parallel with a greater than two-fold increase in serum active GLP-1. Adequately powered, longer-duration trials with pre-specified metabolic endpoints, standardized mixed-meal GLP-1 testing, and enrollment enriched for insulin resistance are needed to further define the scope of metabolic impact and clinical utility of the resM^®^ postbiotic.

## Supporting information

Supplementary Material: Questionnaires

## Declarations

### Author contributions

All authors contributed to study design, data acquisition or analysis, interpretation, and manuscript preparation, and approved the final version.

### Funding

This work was supported by funding from Resbiotic Nutrition, Inc (Dallas, TX, USA), and completed independently by Able Biolabs (Dallas. TX, USA) (Contract Research Organization)

### Conflicts of interest

Some authors are affiliates of Resbiotic Nutrition. The study design and data interpretation were completed independently by Able Biolabs; results are reported in full, including null and unfavorable findings.

### Ethics approval and consent

The study was conducted in accordance with the Declaration of Helsinki and was approved by Sterling Institutional Review Board (Sterling IRB) (Protocol No. RESM101; IRB ID: [13345]). Written informed consent was obtained from all participants prior to study enrollment.

### Trial registration

ClinicalTrials.gov, NCT06911073, registered April 4, 2025 (https://clinicaltrials.gov/study/NCT06911073).

### Protocol and statistical analysis plan

The full study protocol is available from the corresponding author upon reasonable request. A separate statistical analysis plan was not prepared, as all prespecified statistical methods were incorporated into the study protocol before database lock and prior to data analysis.

### Data availability

De-identified individual participant data supporting the findings of this study, together with the study protocol and data dictionary, will be made available by the corresponding author upon reasonable request, subject to review and approval by the sponsor and execution of an appropriate data use agreement. Statistical analysis code is available upon reasonable request. Data will be available beginning upon publication and for five years thereafter.

### Reporting

This manuscript follows the CONSORT 2025 statement.(https://www.equator-network.org/reporting-guidelines/consort/)

## Notes

### Competing Interest Statement

Resbiotic Nutrition, Inc. provided financial and/or in-kind support for this study, and Able Biolabs provided contract research services. Certain authors are employees, consultants, shareholders, officers, or inventors affiliated with Resbiotic Nutrition, Inc. and/or Able Biolabs, including interests related to resM, RSB11, or the studied formulation. All other authors declare no competing interests.

### Clinical Trial

NCT06911073

### Clinical Protocols

https://clinicaltrials.gov/study/NCT06911073

### Author Declarations

Ethics committee/IRB of Sterling Institutional Review Board gave ethical approval for this work.

### Summary of Updates

Correction of formatting issues introduced during the preprint production process. No changes have been made to the scientific content, data, analyses, results, or conclusions.

