## Supplementary Material: Questionnaires for "An Oral Heat-Inactivated Postbiotic Increases Endogenous GLP-1 and Promotes Weight Loss in Adults with Overweight or Obesity: A Randomized Placebo-Controlled Trial"

**Weight**

***Measure your weight in pounds (lb) using the scale you have available. Use the same scale for all weight measurements that you take during the study.***

Enter your weight (lb):

##### **FCQ-T-r questionnaire** **(Cravings Questionnaire)**

A craving is an intense desire for a food. Think about your relationship with food recently and how cravings play a role.

###### **1. When I crave something, I know I won't be able to stop eating once I start.**

Strongly disagree

Disagree

Neutral

Agree

Strongly agree

###### **2. If I eat what I am craving, I often lose control and eat too much.**

Strongly disagree

Disagree

Neutral

Agree

Strongly agree

###### **3. Food cravings invariably make me think of ways to get what I want to eat.**

Strongly disagree

Disagree

Neutral

Agree

Strongly agree

###### **4. I feel like I have food on my mind all the time.**

Strongly disagree

Disagree

Neutral

Agree

Strongly agree

###### **5. I find myself preoccupied with food.**

Strongly disagree

Disagree

Neutral

Agree

Strongly agree

###### **6. Whenever I have cravings, I find myself making plans to eat.**

Strongly disagree

Disagree

Neutral

Agree

Strongly agree

###### **7. I crave foods when I feel bored, angry, or sad.**

Strongly disagree

Disagree

Neutral

Agree

Strongly agree

###### **8. I have no will power to resist my food crave.**

Strongly disagree

Disagree

Neutral

Agree

Strongly agree

###### **9. Once I start eating, I have trouble stopping.**

Strongly disagree

Disagree

Neutral

Agree

Strongly agree

###### **10. I can't stop thinking about eating no matter how hard I try.**

Strongly disagree

Disagree

Neutral

Agree

Strongly agree

###### **11. If I give in to a food craving, all control is lost.**

Strongly disagree

Disagree

Neutral

Agree

Strongly agree

###### **12. Whenever I have a food craving, I keep on thinking about eating until I actually eat the food.**

Strongly disagree

Disagree

Neutral

Agree

Strongly agree

###### **13. If I am craving something, thoughts of eating it consume me.**

Strongly disagree

Disagree

Neutral

Agree

Strongly agree

###### **14. My emotions often make me want to eat.**

Strongly disagree

Disagree

Neutral

Agree

Strongly agree

###### **15. It is hard for me to resist the temptation to eat appetizing foods that are in my reach.**

Strongly disagree

Disagree

Neutral

Agree

Strongly agree

**Scoring instructions:**

Items are scored on a scale from 1 to 6:

- Strongly disagree = 1
- Disagree = 2
- Neutral = 3
- Agree = 4
- Strongly agree = 5

Raw scores are calculated by the sum of all scores for the subscale or total. No additional weights are assigned to the questions to calculate scoring. Total score will range from 15 to 90.

**PHQ9 Questionnaire**

Over the last 2 weeks, how often have you been bothered by any of the following problems?

***1. Little interest or pleasure in doing things***

Not at all

Several days

More than half the days

Nearly every day

***2. Feeling down, depressed, or hopeless***

Not at all

Several days

More than half the days

Nearly every day

***3. Trouble falling or staying asleep, or sleeping too much***

Not at all

Several days

More than half the days

Nearly every day

***4. Feeling tired or having little energy***

Not at all

Several days

More than half the days

Nearly every day

***5. Poor appetite or overeating***

Not at all

Several days

More than half the days

Nearly every day

***6. Feeling bad about yourself or that you are a failure or have let yourself down***

Not at all

Several days

More than half the days

Nearly every day

***7. Trouble concentrating on things, such as reading the newspaper or watching TV***

Not at all

Several days

More than half the days

Nearly every day

***8. Moving or speaking so slowly that other people have noticed, or the opposite, being so restless*** that you are moving around a lot more than usual

Not at all

Several days

More than half the days

Nearly every day

***9. Thoughts that you would be better off dead, or of hurting yourself***

Not at all

Several days

More than half the days

Nearly every day

**Scoring instructions:**

To calculate PHQ-9 scores, items are scored on a scale from 0 to 3 and summed:

- Not at all = 0
- Several days = 1
- More than half the days = 2
- Nearly every day = 3

To interpret total score:

| **Total Score** | **Depression Severity** |
| --- | --- |
| 1-4 | Minimal depression |
| 5-9 | Mild depression |
| 10-14 | Moderate depression |
| 15-19 | Moderately severe depression |
| 20-27 | Severe depression |

**Daily Compliance & AE Check-In**

***1. Have you taken your dose today?***

Yes

No

***2. Do you have any adverse events to report?***

If yes, please list below with the date of the event:

***3. Do you have any changes in medications, supplements, or vaccines to report?***

If yes, please list below with the date of the change:
